# Natural History of Fibrotic Interstitial Lung Disease using AI-driven Test-free Assessment of Routine EHR

**DOI:** 10.64898/2026.08.19.26360827

**Authors:** Dmytro Onishchenko, Fernando Martinez, Anthony N. Gerber, Edward Cantu, Girish Nair, Ishanu Chattopadhyay

## Abstract

**Rationale:** Fibrosing interstitial lung diseases (ILDs), including idiopathic pulmonary fibrosis (IPF), have heterogeneous postdiagnosis courses. Prognostic tools often rely on pulmonary function testing, imaging, or laboratory data that may not be uniformly available, and rarely provide individualized, time-updated forecasts of multiple trajectory events.

**Objectives:** Can longitudinal healthcare claims generate test-free, time-updated forecasts of clinically actionable postdiagnosis trajectory events in patients with fibrosing ILD and IPF?

**Methods:** Using de-identified longitudinal administrative claims from the Merative™ MarketScan^®^ Commercial Claims and Encounters and Medicare Supplemental and Coordination of Benefits databases, we constructed code-based digital twins (ZeBRA) encoding each patient’s evolving diagnosis, pharmacy, and procedure history. Horizon-specific models forecast seven claims-observable events: supplemental oxygen escalation, pulmonary hypertension, acute respiratory failure/ARDS composite, nausea, diarrhea, liver injury, and gastrointestinal bleeding. The analytic cohort included 345,918 patients with fibrosing ILD, including 17,284 with IPF. Predictions were evaluated in a time-updated follow-up setting over 1-month, 6-month, and 1-year horizons.

**Results:** Predictive discrimination was consistent across events and horizons. In fibrosing ILD, AUC ranged from 0.691 for liver injury at 1 year to 0.912 for oxygen dependence at 1 month, with PPV ranging from 0.189 to 0.714. At 1 month, oxygen dependence achieved AUC 0.912 ± 0.005 with PPV 0.473 ± 0.005, and pulmonary hypertension achieved AUC 0.881 ± 0.005 with PPV 0.539 ± 0.005. The IPF subcohort showed analogous horizon-dependent performance, with AUC ranging from 0.687 to 0.855 and PPV from 0.245 to 0.817. At 1 month in IPF, PPV was 0.753 ± 0.015 for oxygen dependence and 0.817 ± 0.011 for pulmonary hypertension.

**Conclusions:** A test-free digital-twin framework from routine longitudinal claims can provide individualized, time-updated forecasts of actionable fibrosing ILD and IPF trajectory events without imaging, pulmonary function tests, laboratory data, clinical notes, or patient-facing data collection. These forecasts may support low-burden reassessment, anticipatory care planning, and earlier recognition of elevated near-term risk for respiratory deterioration or management-altering complications.

## Introduction

Fibrosing interstitial lung diseases (ILDs) comprise a heterogeneous set of disorders characterized by progressive parenchymal remodeling, irreversible loss of lung function, and substantial morbidity and mortality. ^1–3^ Idiopathic pulmonary fibrosis (IPF), a prototypical and common subtype within this spectrum, has informed much of the clinical and therapeutic landscape, but the broader category of fibrosing ILD shares a central clinical challenge: individual patients follow markedly different post-diagnosis courses despite superficially similar baseline presentations. Some individuals remain stable for prolonged periods, while others experience accelerated functional decline, oxygen dependence, pulmonary hypertension, and acute exacerbation. This heterogeneity reflects not only lung-specific injury and repair processes, but also systemic comorbidity burden and evolving patterns of health-care utilization, which together shape the real-world clinical course after diagnosis. ^4^ Existing prognostic approaches typically rely on static baseline measurements such as pulmonary function, imaging features, or composite indices, and often target a single outcome, with limited guidance for anticipating the sequence of clinically meaningful events that unfold over time, including shifting complication risks, escalation of care needs, and transitions to advanced interventions. ^5–7^ Consequently, clinicians face persistent uncertainty in planning surveillance intensity, timing referrals (including for lung transplantation evaluation), and counseling patients about likely near- and intermediate-term milestones. Therapeutic interventions introduce additional complexity; antifibrotics such as pirfenidone and nintedanib can slow functional decline in selected populations but do not halt progression, and real-world patterns of initiation, dose adjustment, intolerance, and discontinuation can themselves become part of the observable post-diagnosis journey. ^8–14^ For many patients with fibrosing ILD, the clinically salient question is therefore not only whether treatment is used, but how evolving comorbidities, complications, and health-system interactions jointly determine outcomes over time.

A patient-specific ability to forecast post-diagnosis natural history—capturing progression milestones, complications, and care transitions across multiple horizons—would represent a fundamental advance for fibrosing ILD. Such forecasting could support personalized surveillance, earlier recognition of destabilization, better-timed referrals, improved trial enrichment, and more informed shared decision-making grounded in expected near-future clinical states rather than static baseline risk.

Here we extend our prior work on AI-enabled point-of-care screening for IPF ^15^ (which demonstrated that routinely collected diagnostic and utilization patterns can identify patients at elevated risk of a future IPF diagnosis without relying on imaging) to the downstream problem of post-diagnosis trajectory forecasting across fibrosing ILD. Building on that screening framework, we infer a personalizable digital twin from real-world electronic health records that models the evolving sequence of medical encounters following diagnosis. Rather than treating outcomes in isolation, our approach represents post-diagnosis course as a dynamic process in which comorbidities, therapeutic exposures, and emerging complications interact over time.

In this study we focus on the subset of clinically meaningful natural-history events that are directly observable and consistently captured in routine longitudinal records, emphasizing progression- and management-defining milestones rather than terminal endpoints. Specifically, we forecast respiratory deterioration markers and downstream care transitions such as escalation to supplemental oxygen and development of pulmonary vascular/right-heart complications, alongside clinically consequential treatment-course events when antifibrotics are used, including gastrointestinal toxicity, liver injury, and bleeding complications. ^16^ Across multiple prediction horizons, the resulting digital twins enable probabilistic forecasting of these outcomes at the individual level, supporting a shift from static post-diagnosis risk stratification toward an individualized, evolving forecast of the patient journey.

## Methods

### Data Source and Cohorts

All predictive models in this study were developed and evaluated using longitudinal administrative claims data from the Merative™ MarketScan® Commercial Claims and Encounters (CCAE) and Medicare (MDCR) databases. These datasets comprise de-identified healthcare utilization records for over 160 million individuals enrolled in employer-sponsored and government health plans across the United States, spanning multiple calendar years. The data include time-stamped diagnostic codes (ICD-9-CM and ICD-10-CM), procedural codes (CPT/HCPCS), and outpatient and retail prescription claims (National Drug Codes), enabling detailed reconstruction of longitudinal clinical trajectories.

Inclusion criteria required continuous enrollment with medical and pharmacy benefits for a minimum lookback period prior to the index date to ensure adequate capture of baseline comorbidity patterns. Patients with insufficient observation windows for post-diagnosis outcome assessment were excluded to prevent temporal bias. Definitions for the analytic disease cohorts and for the seven post-diagnosis target events were based on prespecified structured code sets; the target-event code definitions are provided in e-Table 1.

The final analytic cohort consisted of n = 345,918 patients meeting all inclusion and exclusion criteria (Figure 1a). Model development and evaluation were performed on non-overlapping patient subcohorts. Baseline demographic and clinical characteristics of the fibrosing ILD cohort and the IPF subcohort are summarized in Table 1. As expected, the IPF subcohort was older and exhibited a higher prevalence of cardiopulmonary comorbidities and antifibrotic exposure relative to the broader fibrosing ILD population. Figure 1b,c shows the observed prevalence of the seven tracked events as a function of time since first ILD/IPF documentation, highlighting distinct temporal patterns across progression markers and treatment-course or complication events.

**TABLE 1.**
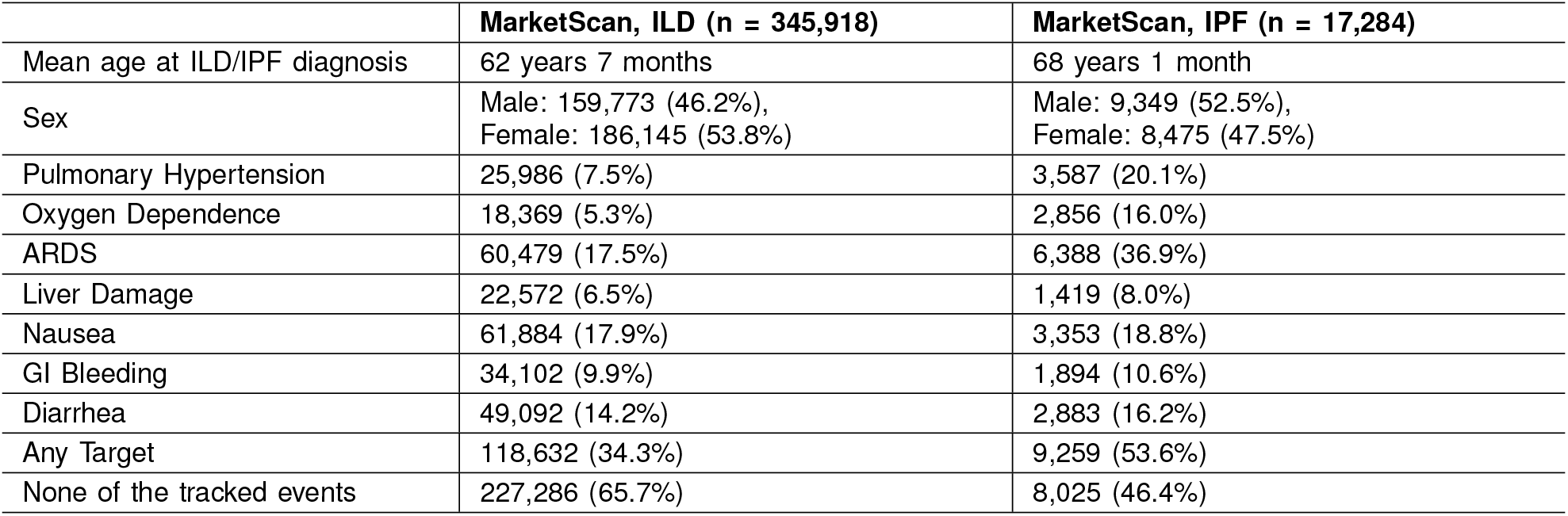
Patient Characteristics.

**Figure 1.**
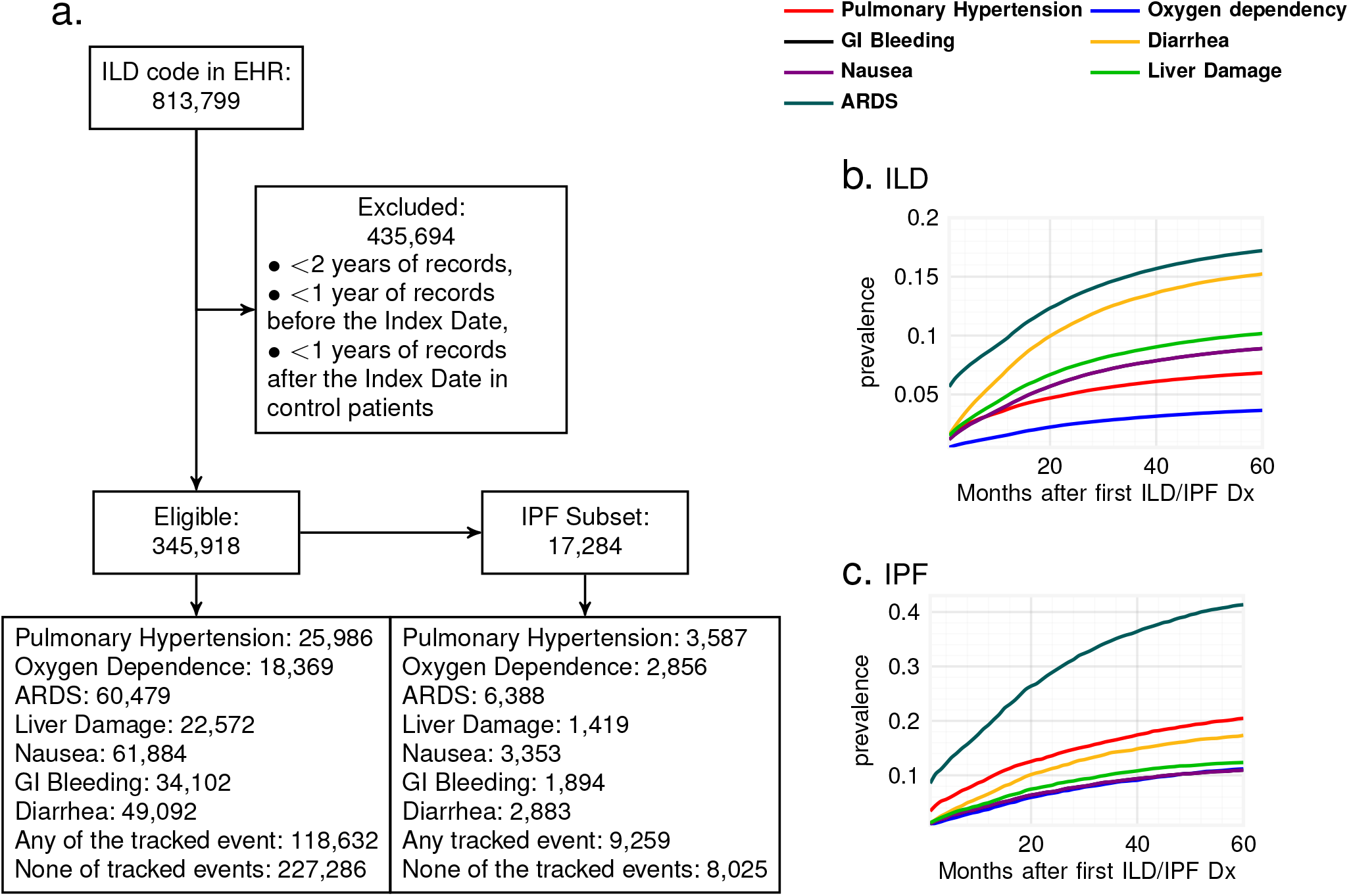
Cohort construction and observed post-diagnosis event prevalence. (a) Cohort selection flow diagram for the fibrosing ILD cohort and the IPF subcohort derived from MarketScan claims data. (b,c) Observed prevalence over time since first ILD/IPF diagnosis for the seven tracked post-diagnosis trajectory events: pulmonary hypertension, oxygen dependence, ARDS, liver damage, nausea, gastrointestinal bleeding, and diarrhea, shown for the fibrosing ILD cohort (b) and the IPF subcohort (c).

### Outcome Definitions and Prediction Settings

We modeled seven clinically meaningful post-diagnosis trajectory outcomes/events that are reliably observable in routine longitudinal records: 1) escalation to supplemental oxygen, 2) development of pulmonary hypertension, and five treatment-course or complication events — 3) nausea, 4) diarrhea, 5) liver injury, 6) development of acute respiratory distress syndrome or respiratory failure (referred to here as ARDS, claims-coded as ICD-10 J80 or J96.x) and 7) gastrointestinal bleeding. These events were selected to reflect progression-defining milestones and management-altering complications rather than terminal endpoints such as hospitalization or mortality. Outcome definitions relied exclusively on structured claims data and were specified a priori using clinically informed code sets (e-Table 1).

Predictions were evaluated under a *time-updated prediction framework* (See Figure 2a), where predictions were generated during follow-up after fibrosing ILD or IPF documentation. For cases, the prediction timepoint was placed 1 month, 6 months, or 1 year before the first occurrence of the target event; for controls, it was placed 12 months before the end of available follow-up. Outcomes were then forecast over forward horizons of 1 month, 6 months, and 1 year from that prediction timepoint.

**Figure 2.**
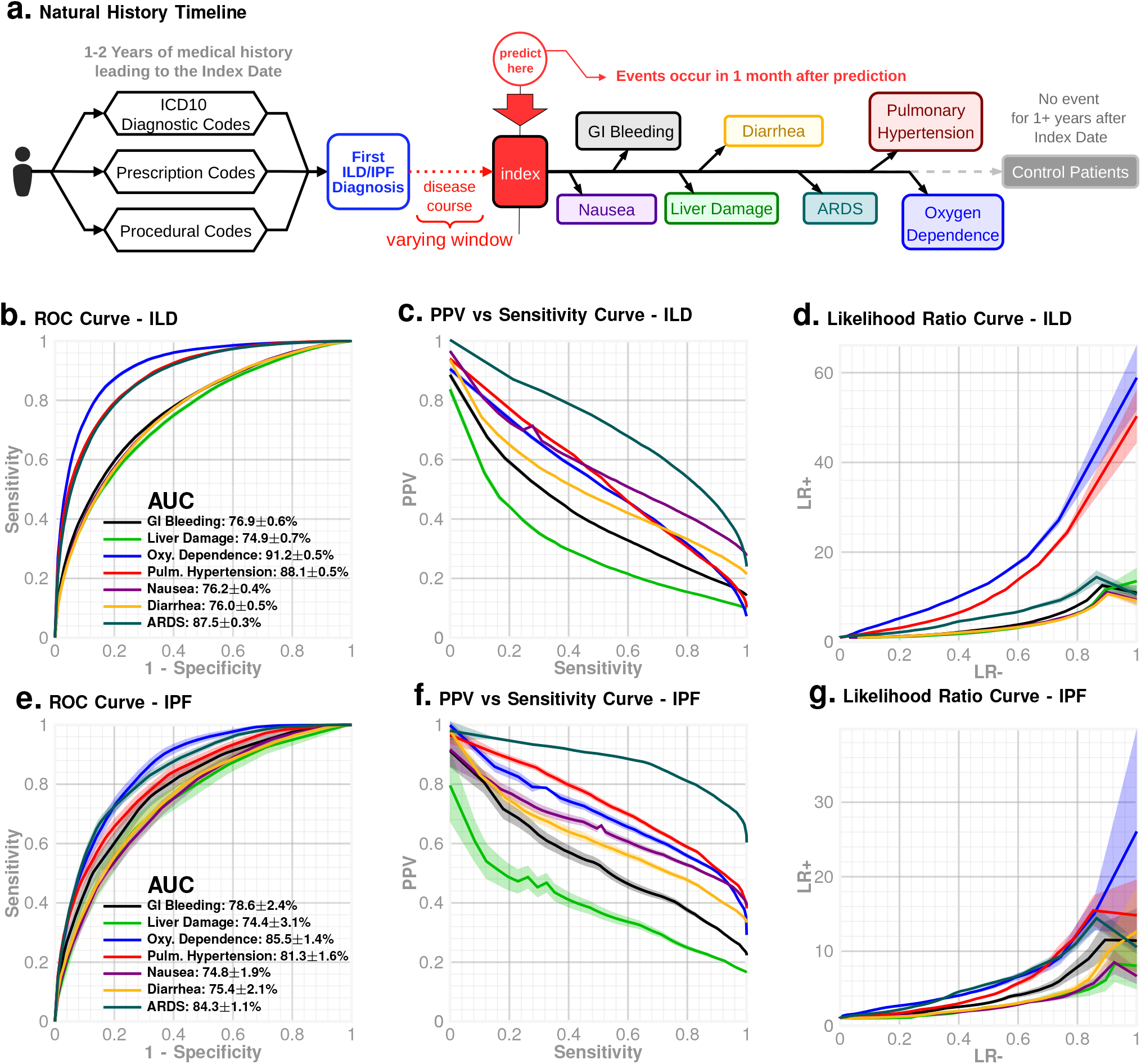
Time-updated prediction framework and 1-month horizon performance. (a) Schematic of the time-updated prediction framework. At any follow-up timepoint after first ILD/IPF diagnosis, risk is estimated from the preceding longitudinal diagnostic, procedural, and prescription history, and outcomes are forecast over the subsequent 1-month horizon. (b–d) Performance in the fibrosing ILD cohort for 1-month-ahead prediction of the seven tracked events, shown as ROC curves (b), PPV versus sensitivity curves (c), and likelihood-ratio curves (d). (e–g) Corresponding 1-month-ahead performance in the IPF subcohort, shown as ROC curves (e), PPV versus sensitivity curves (f), and likelihood-ratio curves (g).

### ZeBRA Framework

Post-diagnosis natural history modeling was performed using the ZeBRA (Zero-Burden Risk Assessment) framework, a purpose-built AI system designed for calibrated risk inference from sparse, noisy longitudinal healthcare records. ZeBRA operates exclusively on structured EHR and claims data, requiring no laboratory values, imaging, pulmonary function tests, or patient-reported inputs.

Each patient is represented as a set of temporally ordered sequences of diagnostic (Dx), medication (Rx), and procedural (Px) codes. These sequences are transformed into engineered features capturing code prevalence, timing, recurrence, and cross-channel dependencies, and into a compact multi-dimensional embedding encoding outcome-specific odds ratios of observed codes. Separate gradient boosting decision tree models implemented as Light Gradient Boosting Machines (LGBM) ^17^ are trained for each data channel (Dx, Rx, Px), and their outputs are combined in a top-layer GBDT ensemble to produce calibrated risk estimates for each outcome (e-Figure 3).

This modular architecture enables robust learning under heterogeneous coding density and variable follow-up, while maintaining interpretability and statistical stability. Model complexity varied by outcome, with trainable parameters ranging from 52,000 to 160,000 depending on the prediction target (e-Table 4).

### Model Training and Evaluation

A total of 21 models were trained (for each of seven prediction targets and each of 3 prediction horizons). Model training and evaluation were conducted using disjoint patient subsets, with no overlap between training and testing cohorts. All features were derived exclusively from data preceding the index date, and no future information leakage was permitted. Predictive performance was assessed using standard discrimination metrics, including area under the receiver operating characteristic curve (AUC), positive predictive value, and likelihood ratios, evaluated on held-out test cohorts. We report overall performance for the PF (ILD) cohort of patients, as well as the subcohort of patients diagnosed with Idiopathic Pulmonary Fibrosis (IPF, ICD-10 code J84.112).

### Ethics and Data Governance

All analyses were conducted on fully de-identified administrative claims data. In accordance with U.S. federal regulations, use of these data does not constitute human subjects research and is exempt from institutional review board oversight.

## Results

### Prediction performance

Validation prediction demonstrated consistent discriminative performance across all seven trajectory events (Table 2, Figure 2). In the fibrosing ILD cohort, AUC ranged from **0.691** (liver damage, 1 year) to **0.912** (oxygen dependence, 1 month), with sensitivity ranging from **0.176** to **0.575** and PPV ranging from **0.189** to **0.714**. LR+ ranged from **3.468** to **11.461**, and LR-ranged from **0.447** to **0.868**. One-month forecasts showed particularly strong performance for oxygen dependence (AUC 0.912 ± 0.005, sensitivity 0.575 ± 0.012, PPV 0.473 ± 0.005, LR+ 11.461 ± 0.237, LR-0.447 ± 0.012), and pulmonary hypertension (AUC 0.881 ± 0.005, sensitivity 0.510 ± 0.010, PPV 0.539 ± 0.005, LR+ 10.085 ± 0.202, LR-0.516 ± 0.011), with gradual attenuation at 6 months and 1 year across outcomes (e-Tables 2 and 3).

**TABLE 2.** Out of sample prediction performance for a horizon of 1 month for studied natural history events^⋆^.

| Event | Cohort | AUC | Sensitivity | PPV | NPV | LR+ | LR- |
| --- | --- | --- | --- | --- | --- | --- | --- |
| Pulmonary Hypertension | ILD | 0.881 ± 0.005 | 0.510 ± 0.010 | 0.539 ± 0.005 | 0.944 ± 0.001 | 10.085 ± 0.202 | 0.516 ± 0.011 |
|  | IPF | 0.813 ± 0.016 | 0.363 ± 0.027 | 0.817 ± 0.011 | 0.708 ± 0.009 | 7.289 ± 0.541 | 0.671 ± 0.028 |
| Oxygen Dependence | ILD | 0.912 ± 0.005 | 0.575 ± 0.012 | 0.473 ± 0.005 | 0.966 ± 0.001 | 11.461 ± 0.237 | 0.447 ± 0.012 |
|  | IPF | 0.855 ± 0.014 | 0.372 ± 0.031 | 0.753 ± 0.015 | 0.786 ± 0.008 | 7.431 ± 0.613 | 0.661 ± 0.032 |
| ARDS | ILD | 0.875 ± 0.003 | 0.477 ± 0.007 | 0.751 ± 0.003 | 0.852 ± 0.002 | 9.452 ± 0.133 | 0.551 ± 0.007 |
|  | IPF | 0.843 ± 0.011 | 0.383 ± 0.020 | 0.921 ± 0.004 | 0.503 ± 0.008 | 7.627 ± 0.407 | 0.649 ± 0.021 |
| Diarrhea | ILD | 0.760 ± 0.005 | 0.272 ± 0.006 | 0.596 ± 0.006 | 0.828 ± 0.001 | 5.383 ± 0.127 | 0.767 ± 0.007 |
|  | IPF | 0.754 ± 0.021 | 0.249 ± 0.027 | 0.711 ± 0.022 | 0.717 ± 0.007 | 5.006 ± 0.537 | 0.791 ± 0.028 |
| GI Bleeding | ILD | 0.769 ± 0.006 | 0.306 ± 0.008 | 0.504 ± 0.007 | 0.892 ± 0.001 | 6.093 ± 0.160 | 0.731 ± 0.008 |
|  | IPF | 0.786 ± 0.024 | 0.298 ± 0.035 | 0.630 ± 0.028 | 0.825 ± 0.007 | 6.227 ± 0.704 | 0.738 ± 0.037 |
| Liver Damage | ILD | 0.749 ± 0.007 | 0.275 ± 0.009 | 0.372 ± 0.008 | 0.924 ± 0.001 | 5.485 ± 0.188 | 0.763 ± 0.010 |
|  | IPF | 0.744 ± 0.031 | 0.229 ± 0.038 | 0.473 ± 0.041 | 0.862 ± 0.006 | 4.731 ± 0.751 | 0.811 ± 0.040 |
| Nausea | ILD | 0.762 ± 0.004 | 0.278 ± 0.006 | 0.714 ± 0.005 | 0.775 ± 0.001 | 5.560 ± 0.114 | 0.760 ± 0.006 |
|  | IPF | 0.748 ± 0.019 | 0.236 ± 0.024 | 0.751 ± 0.019 | 0.659 ± 0.007 | 4.757 ± 0.487 | 0.804 ± 0.026 |

★ LR+: positive likelihood ratio, LR-: negative likelihood ratio, PPV: positive predictive value, NPV: negative predictive value.

The IPF subcohort demonstrated analogous trends, with AUC ranged from **0.687** (nausea, 1 year) to **0.855** (oxygen dependence, 1 month). Across targets and horizons, sensitivity ranged from **0.141** to **0.372**, PPV ranged from **0.245** to **0.817**, LR+ ranged from **3.085** to **7.431**, and LR-ranged from **0.661** to **0.902**. At 1 month, oxygen dependence (AUC 0.855 ± 0.014, sensitivity 0.372 ± 0.031, PPV 0.753 ± 0.015, LR+ 7.431 ± 0.613, LR-0.661 ± 0.032) and pulmonary hypertension (AUC 0.813 ± 0.016, sensitivity 0.363 ± 0.027, PPV 0.817 ± 0.011, LR+ 7.289 ± 0.541, LR-0.671 ± 0.028) again showed the strongest operating characteristics, while adverse-event endpoints remained predictively tractable with clinically meaningful PPV at short horizons.

### Horizon-dependent trends and event-specific behavior

Across both cohorts, prediction performance exhibited consistent horizon-dependent patterns. Discrimination was highest at 1 month, declined at 6 months, and decreased further at 1 year across targets (Tables 2, e-Tables 2, and 3). In particular, short-horizon PPV was substantially higher across outcomes (e.g., ILD oxygen dependence PPV 0.473 ±0.005 at 1 month vs. 0.286 ± 0.007 at 1 year; ILD pulmonary hypertension PPV 0.539 ± 0.005 at 1 month vs. 0.340 ± 0.007 at 1 year; Table 2), reflecting the benefit of conditioning on an updated prediction timepoint in the evolving trajectory.

Collectively, these results demonstrate that digital-twin representations derived from routine longitudinal health records can accurately forecast multiple clinically actionable aspects of fibrosing ILD natural history, enabling higher-precision, near-term forecasting as patient state evolves over time.

Across outcomes, the digital twin models demonstrated strong to acceptable discriminative performance (Figure 2b,e; Table 2). For the 1 month prediction setting, ILD AUCs were: GI bleeding 0.769 ± 0.006, liver damage 0.749 ± 0.007, oxygen dependence 0.912 ± 0.005, pulmonary hypertension 0.881 ± 0.005, nausea 0.762 ± 0.004, diarrhea 0.760 ± 0.005, and ARDS 0.875 ± 0.003.

In the corresponding IPF subcohort, 1 month AUCs were: GI bleeding 0.786±0.024, liver damage 0.744±0.031, oxygen dependence 0.855 ± 0.014, pulmonary hypertension 0.813 ± 0.016, nausea 0.748 ± 0.019, diarrhea 0.754 ± 0.021, and ARDS 0.843 ± 0.011. Importantly, these results are obtained using only longitudinal diagnostic histories available in routine electronic health records, without reliance on pulmonary function testing, imaging, laboratory values, or patient-reported symptoms. This zero-burden design enables consistent performance across heterogeneous care settings and incomplete longitudinal follow-up.

### High-specificity Operating Characteristics

To evaluate clinical utility under realistic deployment constraints, we examined positive predictive value (PPV) as a function of sensitivity (Figure 2c,f). In the 1 month prediction setting for ILD, representative operating points include PPV 0.473±0.005 at sensitivity 0.575±0.012 for oxygen dependence, and ILD pulmonary hypertension PPV 0.539±0.005 at sensitivity 0.510±0.010 (Table 2), consistent with deployment scenarios prioritizing high specificity to minimize false positives.

Likelihood ratio analysis further supports the practical value of the inferred risk scores (Figure 2d,g). In the 1 month time-updated ILD setting LR+ ranged from 5.383 ± 0.127 (diarrhea) to 11.461 ± 0.237 (oxygen dependence) (Table 2). These results indicate substantial post-test risk enrichment for flagged patients and suggest that the digital twin outputs can be used not only for risk stratification, but also to meaningfully alter pre-test to post-test probabilities in clinical decision-making.

## Discussion

This study demonstrates that zero-burden, claims-derived digital twins can be used to forecast clinically meaningful components of the post-diagnosis natural history of fibrosing interstitial lung disease (ILD), including the prototypical idiopathic pulmonary fibrosis (IPF) subset, using only routinely documented diagnostic, procedural, and prescription information. Focusing on seven progression- and management-defining trajectory events that are consistently observable in structured records, we show that longitudinal utilization and comorbidity patterns encode actionable information about how individual patient journeys unfold after diagnosis. *Importantly, the present work is intentionally scoped to these reliably captured natural-history events rather than terminal or system-dependent endpoints such as hospitalization or mortality, which are not modeled here*.

The central clinical framing of the present analysis is time-updated forecasting during follow-up, where risk is re-estimated as the patient evolves and the relevant clinical question is what is likely to happen over the next 1 month, 6 months, or 1 year from the current observed state. In this formulation, the models remain informative across all three horizons, but with a clear and clinically intuitive gradient: performance is strongest for near-term forecasting, remains useful at 6 months, and attenuates further by 1 year as the forecast moves farther from the observed disease state. In the ILD cohort, overall discrimination across outcomes and horizons spans AUC 0.691–0.912, with PPV ranging from 0.189 to 0.714. The strongest 1 month operating characteristics are seen for oxygen dependence (AUC 0.912±0.005, PPV 0.473±0.005, LR+ 11.461±0.237) and pulmonary hypertension (AUC 0.881±0.005, PPV 0.539±0.005, LR+ 10.085±0.202). By 1 year, these same progression-defining endpoints remain tractable but are predictably attenuated, with oxygen dependence at AUC 0.863±0.008 and PPV 0.286±0.007, and pulmonary hypertension at AUC 0.847±0.007 and PPV 0.340±0.007 (Table 2,e-Tables 2,3; e-Figure 1,2). The IPF subcohort shows the same horizon-dependent ordering, supporting the robustness of the time-updated formulation in a clinically more specific but smaller population. Across all outcomes and horizons, IPF performance spans AUC 0.687–0.855 and PPV 0.245–0.817. At 1 month, oxygen dependence (AUC 0.855±0.014, PPV 0.753±0.015) and pulmonary hypertension (AUC 0.813±0.016, PPV 0.817±0.011) again provide the strongest near-term operating characteristics, while at 1 year these outcomes remain predictable with more moderate performance: oxygen dependence reaches AUC 0.784±0.027 with PPV 0.535±0.037, and pulmonary hypertension reaches AUC 0.756±0.024 with PPV 0.606±0.033. The 6 month results fall between these short- and longer-horizon extremes, indicating that the inferred digital-twin state captures not only imminent risk but also meaningful medium-term trajectory information. Taken together, these results argue that time-updated follow-up forecasting, rather than a one-time baseline estimate, is the more natural deployment model for this framework in fibrosing ILD care.

The set of predicted events in this work also motivates a clinically relevant notion of “journey phenotypes” that is compatible with real-world data availability. Rather than predicting a single composite endpoint, the ZeBRA digital twin forecasts multiple, distinct trajectory events that plausibly co-occur, cluster, or trade off in individual patients. The joint predictability of progression markers (oxygen dependence, pulmonary hypertension) and complication/toxicity outcomes (GI symptoms, liver injury, bleeding) suggests that longitudinal comorbidity patterns encode latent patient state that can manifest both as advancing pulmonary disease and as susceptibility to adverse clinical courses. This provides a quantitative basis for stratifying patients into longitudinal phenotypes characterized by differential risk for rapid respiratory deterioration, evolving pulmonary vascular disease, and clinically meaningful complications that may influence treatment persistence and overall morbidity. Such stratification can support individualized surveillance intensity, earlier identification of destabilization, and trial enrichment strategies that target specific trajectory risks rather than broad diagnostic categories.

A distinguishing feature of the approach is its strict “test-free” design. All predictions are generated from structured claims/EHR codes (Dx/Rx/Px), without reliance on pulmonary function testing, imaging, laboratory values, or patient-reported symptoms. This matters operationally because real-world ILD care is often fragmented and longitudinal follow-up is incomplete, particularly outside specialty referral centers. The zero-burden design increases the likelihood of stable performance in heterogeneous care settings and provides a pragmatic foundation for scalable implementation. In addition, because the framework is built as modular ensembles over Dx/Rx/Px channels and a compact odds-ratio embedding, it achieves competitive performance without requiring extremely large parameter counts, which may improve robustness in sparse and weakly labeled settings.

Several limitations should be considered when interpreting these findings. First, event definitions are based on claims codes rather than adjudicated clinical outcomes, which introduces potential misclassification and variation in coding practices across institutions and payers. ^18,19^ This limitation is partially mitigated by the large sample size and by focusing on outcomes with clear coding anchors (e-Table 1), but it remains a key consideration for clinical translation. Second, while the models capture longitudinal correlations in coded histories, they do not distinguish causal pathways; in particular, treatment-course events (e.g., nausea, diarrhea, liver injury, bleeding) should be interpreted as forecasted occurrences in routine records rather than as definitively attributable adverse drug reactions. Third, by design, the present paper does not model hospitalization, mortality, acute exacerbations, or imaging-anchored progression metrics; extending the framework to those endpoints will require either additional data modalities or careful endpoint harmonization across data sources. Fourth, despite careful forward-looking construction, residual confounding and differential follow-up intensity can influence apparent predictability, especially in time-updated analyses where clinical contact patterns may increase near impending events.

These limitations point to natural directions for future work. External validation across independent datasets and health systems will be essential to characterize generalizability and calibration. Incorporating additional modalities such as pulmonary function tests, imaging-derived features, and laboratory markers, when available, could further refine individual trajectory forecasts and enable modeling of endpoints that are not reliably captured by claims alone. Finally, translating the forecasting outputs into usable clinical decision support will require prospective evaluation of actionability: identifying which thresholds and deployment settings maximize benefit while minimizing alert fatigue, and determining how multi-outcome forecasts can be integrated into longitudinal care pathways for fibrosing ILD.

## Conclusion

In summary, we show that ZeBRA-based digital twins can reconstruct and forecast multiple components of post-diagnosis fibrosing ILD natural history using only routine longitudinal healthcare records. By supporting time-updated forecasts across clinically meaningful trajectory events and across 1 month, 6 month, and 1 year horizons, this work shifts prognostic assessment from static, single-endpoint risk estimates toward individualized, evolving journey prediction that is scalable, test-free, and compatible with real-world care.

## Data Availability

A functioning API implementation of the full ZeBRA pulmonary hypertension prediction pipeline, including model input
processing and risk-score generation is available at https://github.com/zeroknowledgediscovery/ZeBRA_nhfild. Source
data used for model training and validation consist of de-identified longitudinal administrative claims and are subject to data-use agreements with their respective data owners; these data cannot be publicly released. Derived summary statistics and scripts sufficient to reproduce the main analyses are available upon reasonable request.

## Acknowledgments

The authors thank the Merative™ MarketScan® Research Databases for providing access to the de-identified longitudinal claims data used in this study. We are grateful to colleagues in pulmonary and data science communities for helpful discussions that informed the framing and interpretation of this work. The project was supported by the NIH National Center for Advancing Translational Sciences through grant number UL1TR001998 for computing resources. The content presented here is solely the responsibility of the authors and does not necessarily represent the official views of the NIH, Merative or any affiliated institutions. The funder had no role in design, execution or reporting of the study.

## Data and Code Availability

A functioning API implementation of the full ZeBRA pulmonary hypertension prediction pipeline, including model input processing and risk-score generation is available at https://github.com/zeroknowledgediscovery/ZeBRA_nhfild. Source data used for model training and validation consist of de-identified longitudinal administrative claims and are subject to data-use agreements with their respective data owners; these data cannot be publicly released. Derived summary statistics and scripts sufficient to reproduce the main analyses are available upon reasonable request.

## Supplementary Information

**e_Table 1.** ICD-10 codes used to define prediction targets.

| Condition | ICD Codes |
| --- | --- |
| ILD | J84.10 J84.111 J84.112 J84.89 J84.9 |
| IPF | J84.112 |
| Liver Damage | K71 K71.0 K71.1 K71.10 K71.11 K71.2 K71.3 K71.4 K71.5 K71.50 K71.51 K71.6 K71.7 K71.8 K71.9 K72 K72.0 K72.00 K72.01 K72.1 K72.10 K72.11 K72.9 K72.90 K72.91 R74.0 R74.01 R74.02 R94.5 |
| Oxygen Dependence | Z99.81 |
| GI Bleeding | K25.0 K25.2 K25.4 K25.6 K26.0 K26.2 K26.4 K26.6 K27.0 K27.2 K27.4 K27.6 K28.0 K28.2 K28.4 K28.6 K57.01 K57.11 K57.13 K57.21 K57.31 K57.33 K57.41 K57.51 K57.53 K57.81 K57.91 K57.93 K92.0 K92.1 K92.2 |
| Pulmonary Hypertension | I27.0 I27.2 I27.20 I27.21 I27.22 I27.23 I27.24 I27.29 |
| ARDS | J80 J96 J96.0 J96.00 J96.01 J96.02 J96.1 J96.10 J96.11 J96.12 J96.2 J96.20 J96.21 J96.22 J96.9 J96.90 J96.91 J96.92 |
| Diarrhea | R19.7 |
| Nausea | R11 R11.0 R11.1 R11.10 R11.11 R11.12 R11.13 R11.14 R11.15 R11.2 |

**e_Table 2.**
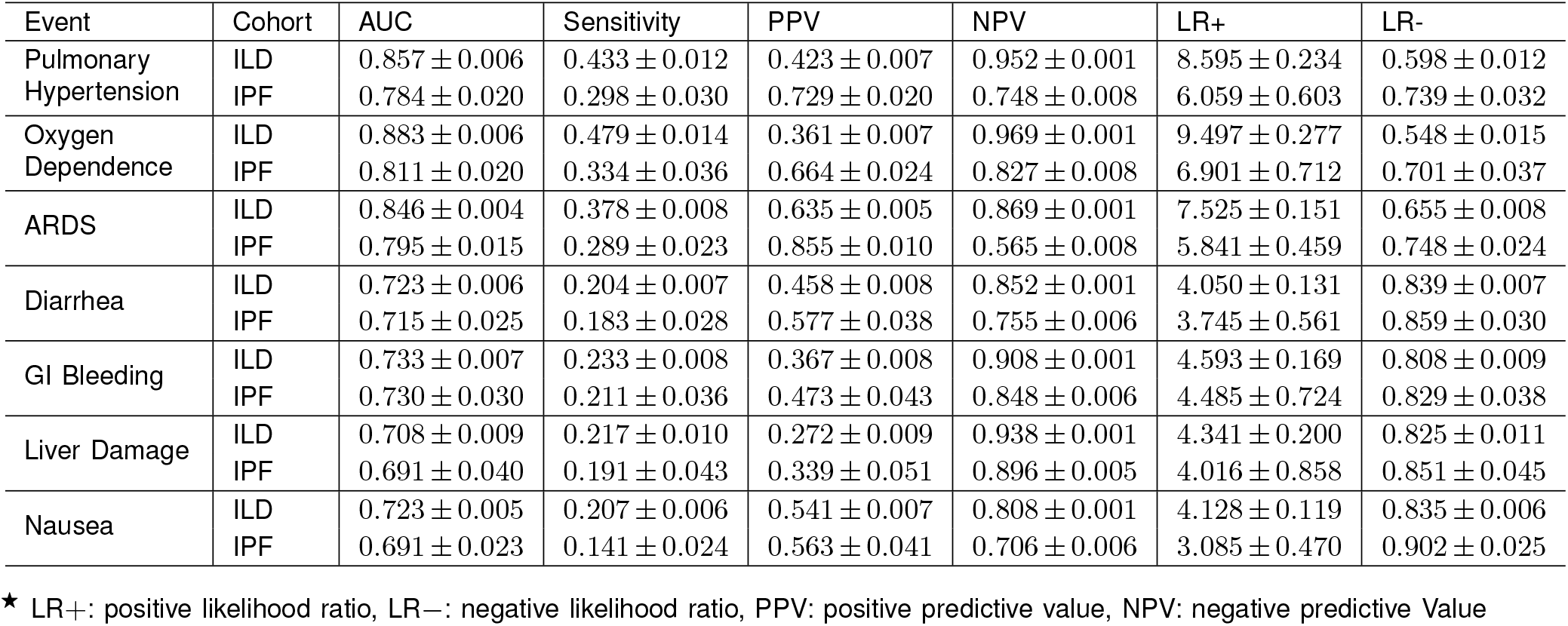
PERFORMANCE TABLE (6 Months Prediction Horizon)^⋆^.

| Event | Cohort | AUC | Sensitivity | PPV | NPV | LR+ | LR- |
| --- | --- | --- | --- | --- | --- | --- | --- |
| Pulmonary Hypertension | ILD | 0.857 ± 0.006 | 0.433 ± 0.012 | 0.423 ± 0.007 | 0.952 ± 0.001 | 8.595 ± 0.234 | 0.598 ± 0.012 |
|  | IPF | 0.784 ± 0.020 | 0.298 ± 0.030 | 0.729 ± 0.020 | 0.748 ± 0.008 | 6.059 ± 0.603 | 0.739 ± 0.032 |
| Oxygen Dependence | ILD | 0.883 ± 0.006 | 0.479 ± 0.014 | 0.361 ± 0.007 | 0.969 ± 0.001 | 9.497 ± 0.277 | 0.548 ± 0.015 |
|  | IPF | 0.811 ± 0.020 | 0.334 ± 0.036 | 0.664 ± 0.024 | 0.827 ± 0.008 | 6.901 ± 0.712 | 0.701 ± 0.037 |
| ARDS | ILD | 0.846 ± 0.004 | 0.378 ± 0.008 | 0.635 ± 0.005 | 0.869 ± 0.001 | 7.525 ± 0.151 | 0.655 ± 0.008 |
|  | IPF | 0.795 ± 0.015 | 0.289 ± 0.023 | 0.855 ± 0.010 | 0.565 ± 0.008 | 5.841 ± 0.459 | 0.748 ± 0.024 |
| Diarrhea | ILD | 0.723 ± 0.006 | 0.204 ± 0.007 | 0.458 ± 0.008 | 0.852 ± 0.001 | 4.050 ± 0.131 | 0.839 ± 0.007 |
|  | IPF | 0.715 ± 0.025 | 0.183 ± 0.028 | 0.577 ± 0.038 | 0.755 ± 0.006 | 3.745 ± 0.561 | 0.859 ± 0.030 |
| GI Bleeding | ILD | 0.733 ± 0.007 | 0.233 ± 0.008 | 0.367 ± 0.008 | 0.908 ± 0.001 | 4.593 ± 0.169 | 0.808 ± 0.009 |
|  | IPF | 0.730 ± 0.030 | 0.211 ± 0.036 | 0.473 ± 0.043 | 0.848 ± 0.006 | 4.485 ± 0.724 | 0.829 ± 0.038 |
| Liver Damage | ILD | 0.708 ± 0.009 | 0.217 ± 0.010 | 0.272 ± 0.009 | 0.938 ± 0.001 | 4.341 ± 0.200 | 0.825 ± 0.011 |
|  | IPF | 0.691 ± 0.040 | 0.191 ± 0.043 | 0.339 ± 0.051 | 0.896 ± 0.005 | 4.016 ± 0.858 | 0.851 ± 0.045 |
| Nausea | ILD | 0.723 ± 0.005 | 0.207 ± 0.006 | 0.541 ± 0.007 | 0.808 ± 0.001 | 4.128 ± 0.119 | 0.835 ± 0.006 |
|  | IPF | 0.691 ± 0.023 | 0.141 ± 0.024 | 0.563 ± 0.041 | 0.706 ± 0.006 | 3.085 ± 0.470 | 0.902 ± 0.025 |

★ LR+: positive likelihood ratio, LR-: negative likelihood ratio, PPV: positive predictive value, NPV: negative predictive Value

**e_Table 3.** PERFORMANCE TABLE (1 Year Prediction Horizon)^⋆^.

| Event | Cohort | AUC | Sensitivity | PPV | NPV | LR+ | LR- |
| --- | --- | --- | --- | --- | --- | --- | --- |
| Pulmonary Hypertension | ILD | 0.847 ± 0.007 | 0.397 ± 0.013 | 0.340 ± 0.007 | 0.960 ± 0.001 | 7.901 ± 0.263 | 0.635 ± 0.014 |
|  | IPF | 0.756 ± 0.024 | 0.229 ± 0.032 | 0.606 ± 0.033 | 0.784 ± 0.007 | 4.835 ± 0.634 | 0.811 ± 0.033 |
| Oxygen Dependence | ILD | 0.863 ± 0.008 | 0.429 ± 0.015 | 0.286 ± 0.007 | 0.973 ± 0.001 | 8.514 ± 0.308 | 0.601 ± 0.016 |
|  | IPF | 0.784 ± 0.027 | 0.280 ± 0.041 | 0.535 ± 0.037 | 0.864 ± 0.007 | 5.663 ± 0.819 | 0.758 ± 0.043 |
| ARDS | ILD | 0.831 ± 0.005 | 0.343 ± 0.008 | 0.547 ± 0.006 | 0.891 ± 0.001 | 6.763 ± 0.168 | 0.692 ± 0.009 |
|  | IPF | 0.778 ± 0.018 | 0.257 ± 0.026 | 0.790 ± 0.017 | 0.637 ± 0.008 | 6.107 ± 0.526 | 0.779 ± 0.028 |
| Diarrhea | ILD | 0.702 ± 0.007 | 0.177 ± 0.007 | 0.361 ± 0.009 | 0.879 ± 0.001 | 3.582 ± 0.142 | 0.866 ± 0.007 |
|  | IPF | 0.689 ± 0.029 | 0.156 ± 0.030 | 0.467 ± 0.049 | 0.798 ± 0.006 | 3.166 ± 0.604 | 0.888 ± 0.032 |
| GI Bleeding | ILD | 0.716 ± 0.008 | 0.207 ± 0.009 | 0.288 ± 0.009 | 0.924 ± 0.001 | 4.096 ± 0.183 | 0.835 ± 0.010 |
|  | IPF | 0.717 ± 0.034 | 0.177 ± 0.039 | 0.359 ± 0.051 | 0.878 ± 0.005 | 3.572 ± 0.784 | 0.866 ± 0.041 |
| Liver Damage | ILD | 0.691 ± 0.010 | 0.188 ± 0.011 | 0.189 ± 0.009 | 0.950 ± 0.001 | 3.969 ± 0.217 | 0.854 ± 0.011 |
|  | IPF | 0.688 ± 0.045 | 0.170 ± 0.048 | 0.245 ± 0.053 | 0.922 ± 0.004 | 3.592 ± 0.961 | 0.873 ± 0.051 |
| Nausea | ILD | 0.699 ± 0.006 | 0.176 ± 0.006 | 0.434 ± 0.009 | 0.841 ± 0.001 | 3.468 ± 0.128 | 0.868 ± 0.007 |
|  | IPF | 0.687 ± 0.028 | 0.164 ± 0.030 | 0.515 ± 0.045 | 0.776 ± 0.006 | 3.296 ± 0.593 | 0.880 ± 0.031 |

★ LR+: positive likelihood ratio, LR-: negative likelihood ratio, PPV: positive predictive value, NPV: negative predictive Value

**e-Figure 1.**
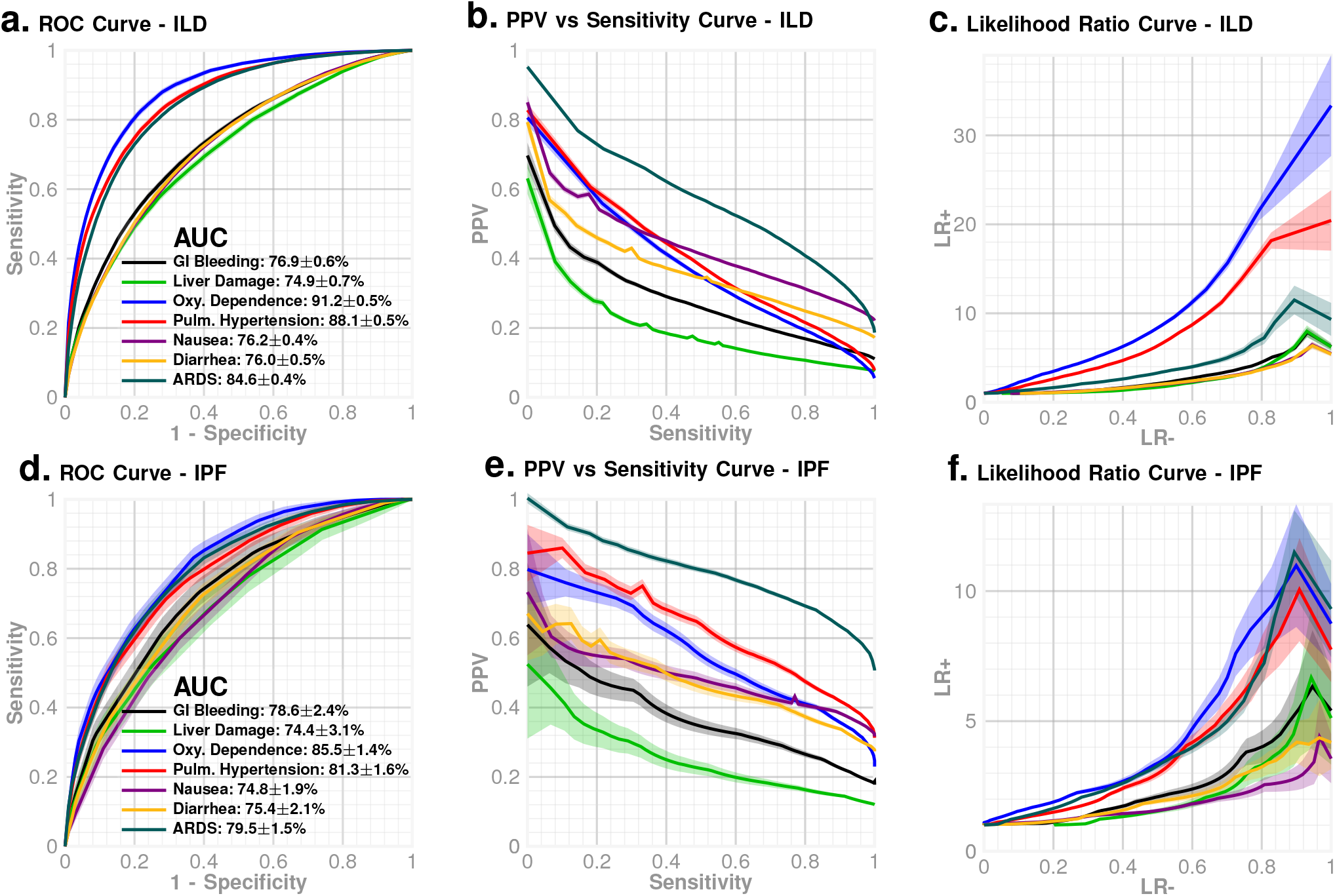
Performance for the 6-month prediction horizon. (a–c) ROC curves, PPV versus sensitivity curves, and likelihood-ratio curves for 6-month-ahead prediction of the seven tracked events in the fibrosing ILD cohort. (d–f) Corresponding 6-month-ahead performance in the IPF subcohort.

**e-Figure 2.**
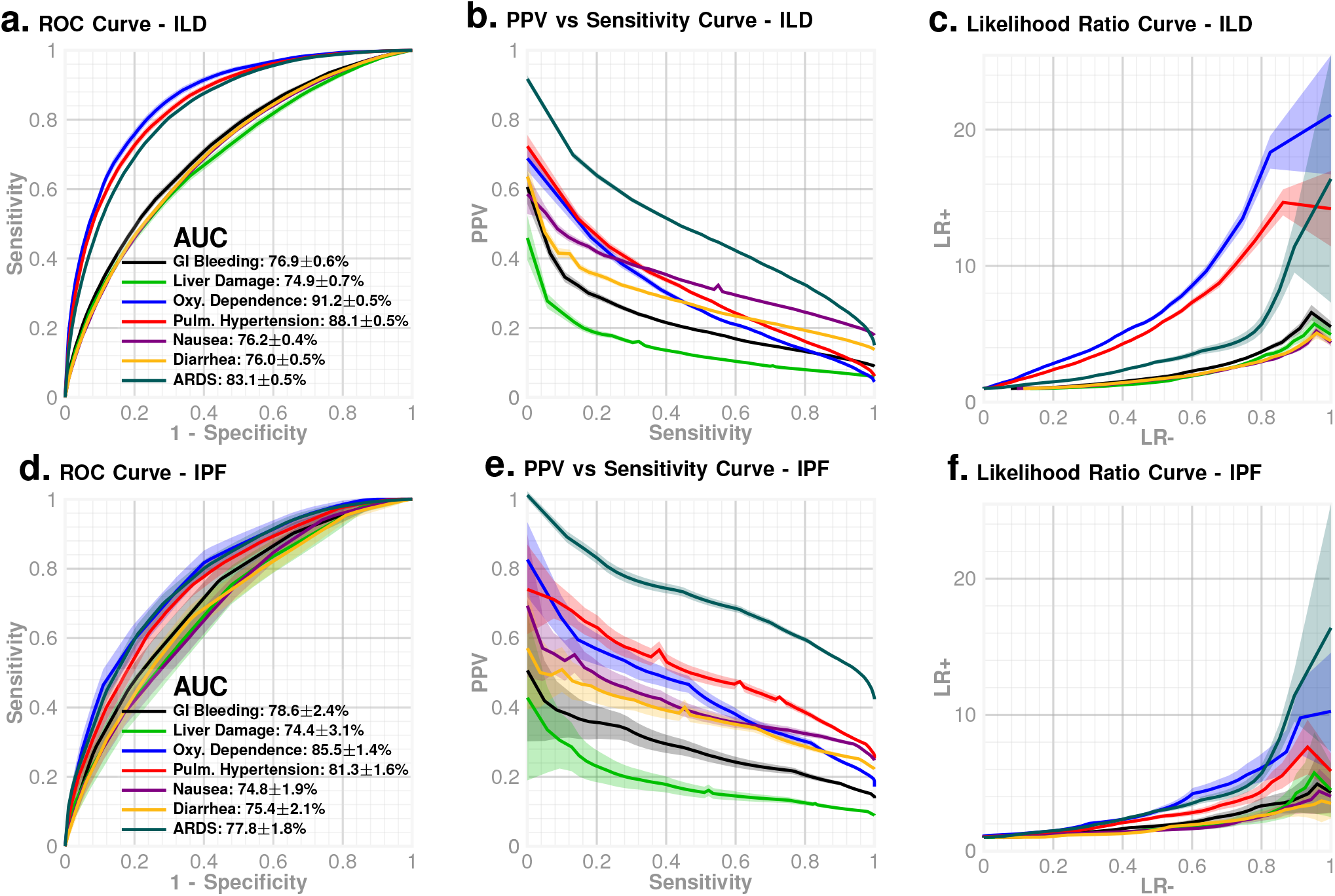
Performance for the 1-year prediction horizon. (a–c) ROC curves, PPV versus sensitivity curves, and likelihood-ratio curves for 1-year-ahead prediction of the seven tracked events in the fibrosing ILD cohort. (d–f) Corresponding 1-year-ahead performance in the IPF subcohort.

## Supplementary Methods: The ZeBRA Algorithm

**e-Figure 3.**
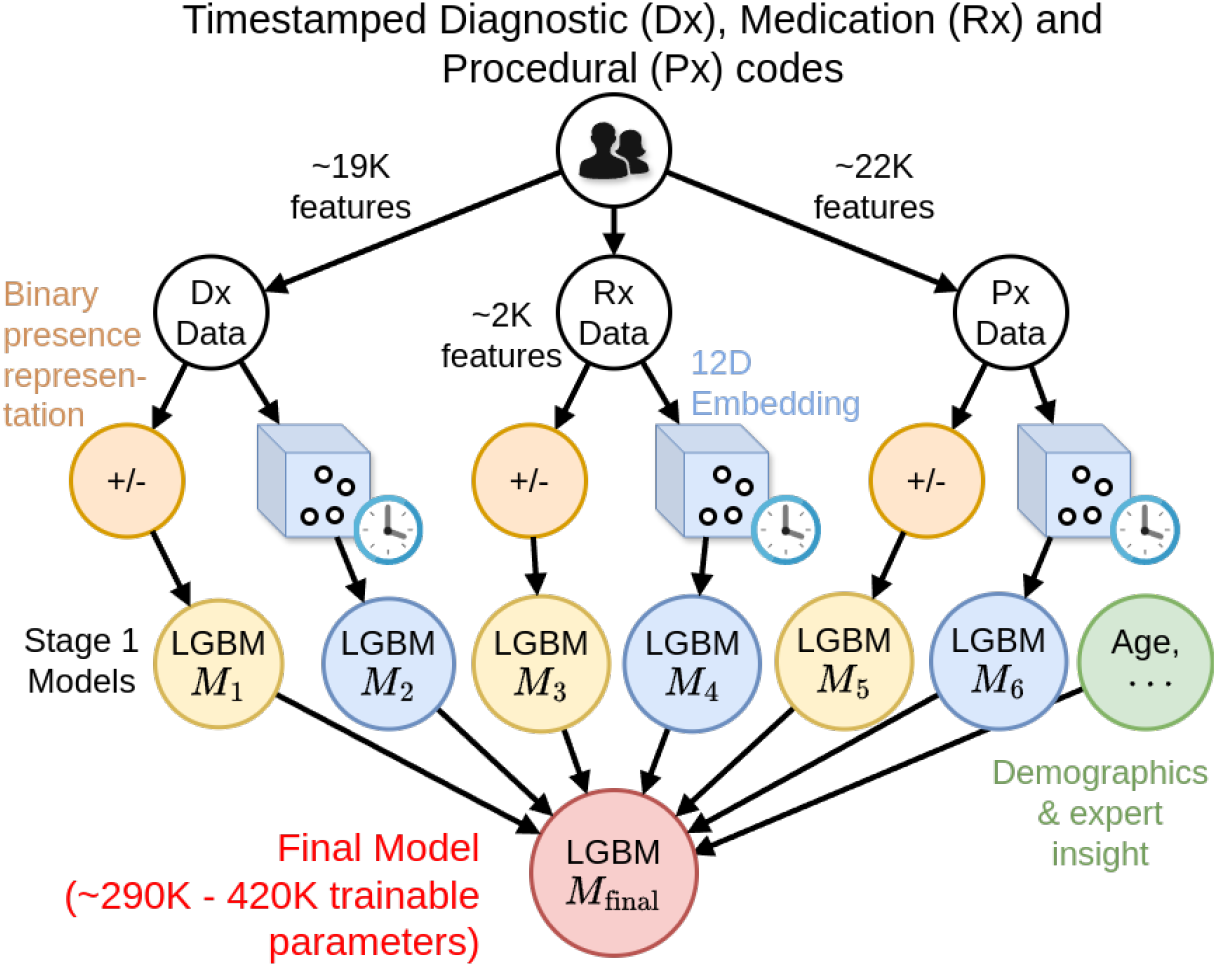
ZeBRA architecture. Combining binary presensce features of over 43K codes with a custom 12 dimensional embedding of Odds ratios of 44K codes given a well-defined target disease phenotype, with a stacked LGBM network, yielding a final model with less than half a million trainable parameters. The emebedding can incorporate longitudinal information, resulting in a scheme that takes advantage of known AI strategies, longitudinal patterns, custom feature engineering, and expert insight into disease pathobiology.

ZeBRA is a general algorithm to predict future clinical documentation of any well-defined target condition or composite endpoint in an individual patient’s Electronic Health Record (EHR), where the target is specified by a pre-defined set of ICD-10 (and related) codes.

### Prediction Task

For each patient *x*, a screening index point *t*_0_ is defined, and the model estimates the risk of a target event; typically the first occurrence. Separate models estimate the risk of occurrence at different prediction horizons, *e*.*g*. 1 week, 1 month or 6 months in future post-*t*_0_.

### Data Source

Training data are drawn from large EHR and claims databases, *e*.*g*. the Merative MarketScan claims dataset ^20^ (with adequate train/validation split), and comprises diagnostic (Dx), procedural (Px), and pharmacy (Rx) codes. The performance results reported for the different scenarios are obtained for out-of-sample held-back data from the database, and are being validated for independent databases such as the All-of-Us Research program ^21^. Patients are stratified by sex, and possibly also by age when it is relevant. ZeBRA is trained on cohorts which exclude patients who do not have a pre-specified length of visibility in the datsbase, typically 1−2 years. Controls are age-matched, target-code-free, with a pre-specified length (typically ≧ 3 years) over which they are visible in the datbase being used.

### Timeline and Observation Window

For cases, the screening index point is set prior to the first target code by the length of the prediction horizon. For controls, the index point is generally 2 years before their final record to ensure target-free follow-up of two years after screening index point. Thus, the observation window (the data used for training for each patient) spans at least 1 year before the index point. These values can be altered to suite a particular problem.

**e_Table 4.** Feature and Parameter Counts BY Target and Prediction Horizon.

| Prediction Target | # DX Features | # DX Trainable Parameters | # RX Features | # RX Trainable Parameters | # PX Features | # PX Trainable Parameters | # Total Trainable Parameters |
| --- | --- | --- | --- | --- | --- | --- | --- |
| Pulmonary Hypertension | 6,374 | 56,158 | 915 | 44,680 | 4,134 | 60,144 | 160,982 |
| Oxygen Dependence | 6,372 | 52,746 | 900 | 50,651 | 4,122 | 54,399 | 157,796 |
| ARDS | 6,296 | 75,764 | 912 | 61,255 | 4,080 | 69,084 | 206,103 |
| Diarrhea | 6,355 | 52,671 | 909 | 29,732 | 4,169 | 44,041 | 126,444 |
| GI Bleeding | 6,376 | 53,324 | 916 | 33,132 | 4,177 | 47,010 | 133,466 |
| Liver Damage | 6,347 | 37,408 | 923 | 24,459 | 4,166 | 37,869 | 99,736 |
| Nausea | 6,278 | 60,048 | 917 | 38,594 | 4,077 | 56,176 | 154,818 |

### Feature Engineering

Denoting the medical history *H*_*j*_(*x*) for each patient *x* is denoted as:

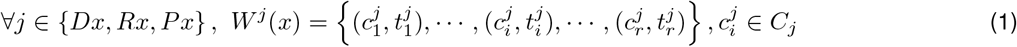

where 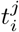 are logging time points for the recorded codes, and *C*_*j*_ are set of valid codes for each channel. We map each patient’s medical history to representations, namely the binary presense feature vector, and the odds-ratio embedding vector described next.

### Prescription Drug Coding

Prescription drug (RX) codes in our primary (National) dataset are provided in the National Drug Code (NDC) format ^22^, which is designed to facilitate the identification and commercial distribution of pharmaceuticals. This coding system, as well as the widely adopted RxNorm ^23^ emphasizes product-specific and manufacturer details of the generic drugs rather than their therapeutic classification and characteristics, thus hindering its use for EHR data analysis.

To facilitate the analysis of prescriptions according to their therapeutic effect, we developed a custom NDC-compatible coding scheme to convert all RX codes in the used datasets. This system, analogous to ICD10 for diagnostic codes, progressively details therapeutic information of a generic drug with each successive character in the code. Each RX code begins with an “rx” prefix to enhance readability and facilitate parsing. This is followed by three alphanumeric characters encoding, respectively, the Therapeutic Group, Therapeutic Class, and Therapeutic Subclass, latter derived from the Therapeutic Class column in the RED BOOK. In cases where any of these attributes are missing or listed as NEC (i.e., not elsewhere classifiable), we use the placeholder character “X”. Finally, to identify a specific generic drug, we append a numeric identifier indicating its order within the set of drugs sharing the same initial five-letter therapeutic code.

### Code Presence Embedding Vectors

For each EHR data channel *j* ∈ *D*, we define a binary presence embedding based on the patient observation window *W* ^*j*^(*x*). Let *C*^*j*^ denote the set of all distinct codes observed across the embedding inference set, augmented with hierarchical code prefixes to enable generalization across varying code granularities.

Specifically, we include for each code *c ∈ C*^*j*^ its prefixes according to the following scheme:

- DX: prefixes of length 1, 2, 3, 5 and 6,
- RX: prefixes of length 3, 4, 5, and 8,
- PROC: prefixes of length 1, 2, 3, 4, and 5.

We define the augmented code set *C*^*j*^ for channel *j* as:

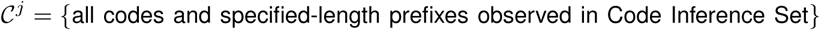

Let *O*^*j*^ (*x*) be the set of codes and code prefixes present in the observation window *W* ^*j*^(*x*) for patient *x*. Then, the presence vector 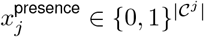 is defined elementwise as:

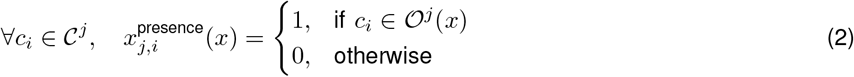

In total, the number of codes and code prefixes we track is *>* 20, 000 for *D*_*X*_ channel, *>* 2000 for *R*_*X*_ channel, and *>* 17, 000 for *P*_*X*_ channel.

### Odds Ratio Embedding Vectors

For each code *c* ∈ *C*^*j*^, in each data channel *j* ∈ *D*, we compute the dictionary of cubed odds ratios based on patients from the Code Inference Set:

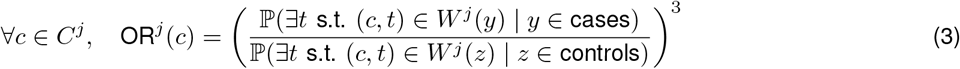

Then we compute odds ratios for all codes in the patients’ observation windows:

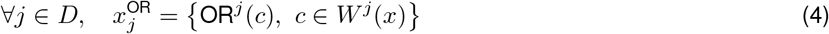

and finally map the computed odds ratios of each patient *x* to a 12-dimensional embedding via aggregation functions

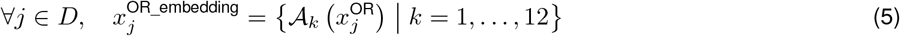

e-Table 5 enumerates for the complete list of aggregation functions *A*_*k*_.

## Model Architecture

ZeBRA is a stacked ensemble of LightGBM models ^17^ with 6 base classifiers (e-Figure 3):

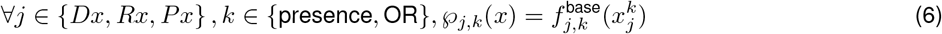

**e_Table 5.** Odds Ratios Aggregations.

| Aggregation Name | Formula |
| --- | --- |
| mean_or | $\text{mean}(r), \forall r \in x_j^{\text{OR}}$ |
| max_or | $\text{max}(r), \forall r \in x_j^{\text{OR}}$ |
| sum_or | $\text{sum}(r), \forall r \in x_j^{\text{OR}}$ |
| std_or | $\text{standard\_deviation}(r), \forall r \in x_j^{\text{OR}}$ |
| range_or | $\text{max}(r) - \text{min}(r), \forall r \in x_j^{\text{OR}}$ |
| geomean_or | $\text{geometric\_mean}([r \text{ if } r \neq 0 \text{ else } 10^{-7}], \forall r \in x_j^{\text{OR}})$ |
| variation_or | $\text{variation}([r \text{ if } r \neq 0 \text{ else } 10], \forall r \in x_j^{\text{OR}})$ |
| meanstd_or | $\text{mean\_or} \times \text{std\_or}$ |
| meanrng_or | $\text{mean\_or} \times \text{range\_or}$ |
| meanstd_sq_or | $\text{mean\_or}^{\text{std\_or}}$ |
| geomeanrng_or | $\text{geomean\_or} \times \text{range\_or}$ |
| geomean_sq_sum | $\sqrt{\text{geomean\_or}^{\text{sum\_or}}}$ |

These six risk outputs are used to train a final LGBM classifier, which also takes as input patient age, and other demographic characteristics (referred to as the set CHAR), to estimate the final ZeBRA risk *℘*:

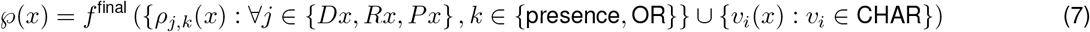

### Training

The set of eligible patients from the database under consideration is split typically 66% - 34% for training and validation. The training set is further split 60/40 for computing odds ratios and training the final LGBM model. We consider almost all unique diagnostic, medication and procedural codes as feature inputs, stratified into the Dx, Rx and Px channels, using in total 43,609 and 42,858 feature inputs respectively for the models corresponding to 50+ (older patients) and 25-50 (younger patients) age groups. The overall ZeBRA model has sufficient capacity for the EHR data at-hand, and we can measure the model complexity as the number of independent (or trainable) parameters (See e-Table 4, number of trainable parameters for the problems targeted here range betwee 35,748 - 68,320).

Gradient boosting machines, which are the conceptual foundations for LighGBMs, are decision-tree ensembles, and the ZeBRA framework, with its stacked network of LGBMs, realizes a non-linear classifier that optimizes the contribution of different EHR data modalities, while making sure the model architecture is not too unconstrained. In particular, ZeBRA avoids off-the-shelf embeddings (*e*.*g*., transformer architectures) in favor of a custom 12D embedding of odd-ratios of Dx, Rx and Px codes. Such neural models use hundreds of millions of parameters (2 orders of magnitude more in this application), ZeBRA’s more limited parameter count (< 0.5M) enables more efficient training, faster inference, better generalization in sparse, weakly labeled data, and robustness to population shifts and health system idiosyncrasies.

## Statistical Analyses, Models and Sufficiency of Sample Size

The ZeBRA performance is measured using standard ML measures such as AUC, likelihood ratios, PPV, NPV and others, as has been demonstrated in several past studies ^15,24,25^. Our sample size needs to be adequate when we train and evaluate ZeBRA. Here the sample size sufficiency question relates to if we would have sufficient number of patients to acceptably bound the uncertainty of our predictions, to confidently validate the tool. The CI for the AUC are calculated using the equivalence with the Mann-Whitney U statistic ^26–28^. We carry out bootstrapped runs over randomly selected sub-cohorts estimating the empirical distribution of AUC over these runs; in the example shown, the mean AUCs obtained by this approach were within ±2% of the U statistic estimate for sufficiently large sub-cohorts. The CI for specificity and sensitivity are computed via the asymptotic method for single proportions ^29^ (the Wald Method). CI for the remaining metrics (PPV, likelihood ratios) are computed from the extremal values of the CI for specificity and sensitivity. We also compute p-values for the null hypotheses that PCoR performance is different from a corresponding baseline for the same sex and dataset using a small fixed set of risk factors (presence or absence of specific mental disorders), using Dantzig’s upper bound on AUC variance ^30,31^. In the example shown, these considerations suggest an estimated minimum sample size of 1435 (1 week horizon) - 1988 (6 month horizon) to yield AUC estimates with a maximum error of < ±0.02 with 95% probability, assuming a 10% target prevalence. Generally, this is a highly realizable goal given the patient numbers we typically encounter in a hospital system.

